# Normative Temperature, Demographic Categories, and the Limits of Infrared Thermometry

**DOI:** 10.64898/2026.10.19.26365925

**Authors:** Anaïs Lohier, Angela G. Campbell, David Chartash

**Affiliations:** Department of Biostatistics, School of Public Health, Yale University, New Haven, CT, USA; Department of Applied Health Science, School of Public Health, Indiana University Bloomington, Bloomington, IN, USA

**Keywords:** infrared thermography, body temperature measurement, measurement uncertainty, non-contact thermometry, subgroup analysis

## Abstract

**Objective:** Temperature readings are routinely stratified by race and ethnicity, though significant group differences may reflect physiology, instrument noise, or categorization rather than biology. This study tests whether these categories partition facial infrared thermography measurements beyond individual variation and baseline device noise.

**Approach:** A secondary analysis of thermography data (*n* = 951 adults) compares inner-canthus maximum (*T*_CEmax_) and full-face maximum (*T*_max_) temperatures across five administrative categories via one-way analysis of variance (ANOVA), mixed-effects modeling, Bland–Altman analysis against oral reference, and Monte Carlo sensitivity analysis.

**Main results:** ANOVA returns significant group differences (*p* < 0.001; *η*^2^ = 0.04 for *T*_CEmax_, 0.03 for *T*_max_), but the total spread (0.2 °C to 0.3 °C) is comparable to device accuracy and within-subject variability. Individual variation accounts for 95% of *T*_CEmax_ variance, and adding simulated Gaussian noise to participant-level temperatures (*σ* = 0.10– 0.30 °C) preserves the complete group-mean rank order in as few as 37.5% of iterations.

**Significance:** Testing whether a group difference survives perturbation at the scale of instrument accuracy offers a check against treating significance alone as evidence of physiology. Here, observed group differences are small relative to measurement variability, and their physiological interpretation remains uncertain, with implications for clinical tools that treat administrative categories as biologically meaningful.

## 1 Introduction

For more than a century, body temperature has been anchored to a nineteenth-century base-line of a 37 °C norm and a 38 °C fever threshold [1] that persists more by convention than biological universality. Non-contact infrared thermography estimates this value at scale using algorithmic modeling, yet its nominal accuracy operates at the same sub-degree scale as typical between-group clinical differences [2, 3]. Despite this narrow margin, these measurements are routinely stratified across race and ethnicity, administrative categories whose origins lie outside the physical measurement process [4, 5].

A statistically significant difference between race and ethnicity groups is straightforward to obtain but difficult to interpret. The single value being tested conflates individual physiology, instrument uncertainty, skin pigmentation and other optical effects, and the administrative encoding of the category itself, yet a standard significance test may not isolate these distinct components. To address this, we introduce a generalizable methodological check for health informatics by testing whether observed demographic group differences survive perturbation at the scale of the instrument’s own accuracy. Applying this framework to a controlled facial infrared thermography dataset of *n* = 951 adults, we evaluate how these encoded categories partition clinical measurements and examine whether statistical significance reflects true physiological divergence or an artifact of measurement noise.

## 2 Background

### 2.1 Standardization and the Construction of Thermal Norms

Clinical thermometry became useful when instruments, anatomical sites, and bedside routines were standardized, making readings comparable across patients [6]. Wunderlich’s nineteenth-century work established 37 °C as a normative mean and 38 °C as a fever threshold, though both were always contingent on instrument and site rather than fixed biological truths. Demographic variation was noted but never systematically addressed, with his brief commentary on population differences relying entirely on a single anecdotal reading borrowed from Livingstone’s *Missionary Travels* [1, 7]. Temperature thus functions as a biometric whose clinical authority is an artifact of standardization, contingent on the instruments, protocols, and historical population that produced it. Contemporary longitudinal analyses confirm that individual baselines vary widely and systematically with age, sex, circadian timing, and body composition [8–10]. Consequently, a rigid threshold derived from a historical population average may fail to reflect individual clinical reality.

### 2.2 Infrared Thermography and Measurement Uncertainty

Infrared thermography is a non-contact method that estimates skin-surface temperature from emitted thermal radiation, and the reported value is a modeled quantity shaped by skin emissivity, calibration state, sensor geometry, ambient conditions, and device-specific radiance-to-temperature conversion algorithms [2, 3, 11]. Skin emissivity, which expresses how closely a surface radiates relative to an ideal blackbody, is typically assumed at approximately 0.98 and is generally stable across skin tones under controlled conditions [12]. Even so, apparent temperature differences across pigmentation levels have been reported in some measurement contexts, while single-point screening has shown no skin-tone effect [13, 14]. Because race and ethnicity categories correlate broadly with skin pigmentation in many populations [15], these measurement effects can map onto demographic labels and produce the appearance of group-level physiological differences without any underlying thermoregulatory basis.

Clinical-grade infrared systems are typically specified at *±*0.3 °C under controlled conditions, reflecting combined contributions from sensor precision, skin emissivity assumptions, ambient stability, and acquisition geometry [3, 16]. This tolerance sits at the same scale as both the inter-individual spread of resting baselines and the group-level differences that demographic stratification typically produces, so the device’s accuracy and the effects under study are the same size [8, 9]. Sustained performance at nominal tolerance requires calibration stability, and infrared sensors are susceptible to drift from optical contamination and component aging, requiring periodic verification against traceable blackbody standards [3]. In practice many non-contact systems fail to meet ISO and ASTM performance criteria even under controlled testing [3, 16]. The large-scale deployment of infrared fever screening during the COVID-19 pandemic showed this failure at scale, as systems installed without blackbody reference targets and under inconsistent facial geometry could not reliably distinguish febrile from afebrile individuals [17].

### 2.3 Demographic Fields as Measurement Metadata

Physiological measurements and demographic fields enter the same analytic frameworks but originate through fundamentally different processes. Temperature values are produced by calibrated sensing systems, while demographic attributes such as race and ethnicity are socially defined, administratively encoded variables reflecting mixtures of self-identification, institutional assignment, and legacy coding [4, 5]. Unlike temperature, these categories originate outside any measurement process and lack independently validated correspondence to physiological states [4, 18].

Once represented as data fields, however, demographic categories are analyzed alongside physiological measurements. Statistical tests then treat observed group differences as a single aggregate effect, collapsing variation that arises separately from physiology, instrumentation, or categorization itself. The present analysis evaluates that combined effect against the empirical accuracy limits of infrared thermometry systems. When this collapsed effect is treated as biological fact, an administrative artifact can be built directly into clinical thresholds and predictive models, giving a category with no physiological basis the same authority as a validated measurement.

## 3 Methods

### 3.1 Data Source and Acquisition Conditions

This study analyzes a publicly available thermographic dataset from PhysioNet, drawn from a controlled clinical evaluation by Zhou *et al*. in which facial thermal images, visible-light images, and oral temperature measurements were collected under Institutional Review Board (IRB) and Food and Drug Administration (FDA)-related study governance [19, 20]. Data collection followed a fixed protocol designed to minimize environmental variability: imaging took place indoors at a university setting, participants acclimated for approximately 15 minutes before measurement, and ambient temperature was held within a narrow range. Camera placement, participant posture, and imaging distance were kept constant throughout, and each participant was imaged multiple times within a single session under identical conditions, yielding repeated measurements of the same thermal state. As this study is a secondary analysis of publicly available, de-identified data, IRB approval was not required.

### 3.2 Variable and Cohort Selection

This analysis focuses on inner-canthus maximum (*T*_CEmax_) and full-face maximum (*T*_max_), defined as the highest radiometric temperature within standardized facial regions of interest (ROIs) following established definitions [20]. These measures have been validated against oral reference temperatures and shown to perform well in fever screening contexts [19, 20].

Facial thermography data were collected using screening systems built to IEC 80601-2-59 and ISO/TR 13154 standards [19], each paired with an external blackbody reference for drift compensation. The system used here (IRT-2; ICI 8640 P-series) is a 640 *×* 512 radiometric long-wave infrared imager (7 μm to 14 μm, 30 Hz) with a manufacturer-specified NETD below 20 mK [21]. While the manufacturer’s uncalibrated standard mode specifies an absolute accuracy of *±*1 °C [21], the system-level accuracy improves to approximately *±*0.2 °C over the skin-temperature range when paired with an external blackbody reference as evaluated in the source study [19, 20], selected for its closer agreement with oral reference temperature and stronger fever-screening performance relative to the alternative system [19, 20]. The *±*0.3 °C specification cited earlier describes clinical-grade non-contact systems as a class and is used only to situate the general scale of device tolerance, whereas all device-specific reasoning here uses the IRT-2’s blackbody-corrected *±*0.2 °C specification. Restricting analysis to a single device limits generalizability across instruments but avoids accumulating calibration uncertainty across systems with differing accuracy margins, a concern that is especially acute when the effects of interest are smaller than nominal device tolerance, and full calibration histories and uncertainty budgets remain unavailable for either system.

To reduce age-related variation in baseline temperature across the lifespan, the cohort was restricted to adults aged 18–30, retaining twelve participants from a broader, overlapping “21– 30” bin since they fell within this range [8, 10]. Race and ethnicity were analyzed across five recorded categories, excluding American Indian or Alaska Native data due to a small sample size (*n* = 3). Neither the PhysioNet release nor its associated publications document how these categories were defined or assigned, including whether they were self-reported by participants or recorded by study staff [19, 20]. In the absence of this documentation, race and ethnicity are treated strictly as administrative labels to examine how this institutional labeling partitions measurements within a controlled thermographic system [4, 5].

### 3.3 Preprocessing and Feature Construction

Thermal images were collected in multiple rounds per participant within a single session. Before averaging, raw round-to-round repeatability was assessed for participants with two or more valid rounds, with the pooled within-subject standard deviation across rounds being 0.14 °C for *T*_CEmax_ and 0.47 °C for *T*_max_. A participant-level mean was computed across all available rounds with no imputation performed. All participants contributed at least one measurement; missingness was 5.47 % in Round 1 and rose to 11.15 % to 13.25 % in Rounds 2 through 4 for both measures. A sensitivity analysis confirmed that the number of available rounds did not significantly modify the ethnicity effect (interaction *p* = 0.065 for *T*_CEmax_ and *p* = 0.101 for *T*_max_).

### 3.4 Statistical Model

Between-group differences in mean facial temperature were evaluated using one-way analysis of variance (ANOVA), specified separately for *T*_CEmax_ and *T*_max_, with the participant’s race and ethnicity as the sole predictor. Covariates were intentionally omitted from this primary ANOVA to evaluate the unadjusted, naive data partitioning typically performed in baseline clinical stratification routines. Model assumptions were assessed using Shapiro–Wilk tests and Brown– Forsythe tests for homogeneity of variance. Kruskal–Wallis tests were run as a nonparametric check. Effect size was quantified using *η*^2^, and significance was evaluated at *α* = 0.001. Pairwise comparisons were evaluated using Tukey HSD at the conventional *α* = 0.05, distinct from the *α* = 0.001 threshold applied to the primary omnibus tests. This ANOVA used one value per participant, computed as each participant’s mean across all available rounds.

A separate linear mixed-effects model used the round-level data directly, retaining every individual round-level observation instead of collapsing to a participant mean. This model was fit to decompose within-versus between-subject variance and to account for repeated measures, specified separately for each outcome using *temp ∼ Ethnicity* + *Gender* + *round* +(1|*SubjectID* ), with random intercepts for each participant. Gender was included as a covariate consistent with reporting guidelines for gender-based analysis [22]. Intraclass correlation coefficients (ICC) were derived from the random-effects variance components.

Agreement between facial temperature and oral reference was assessed using Bland-Altman analysis [23], computed at the participant level (*n* = 951) using the same participant-level means as the ANOVA. Bias and 95% limits of agreement (LoA) were calculated for each outcome, and group-level bias was examined across ethnicity categories to characterize systematic variation in facial–oral discordance. Oral temperature is used as the reference because it is the standard against which *T*_CEmax_ and *T*_max_ were originally validated for fever screening [19, 20]. Oral thermometry is not error-free either; the source study reports an oral thermometer accuracy of

*±*0.1 °C, confirmed against a NIST-traceable reference [19]. The bias and LoA reported below therefore reflect combined facial and oral measurement uncertainty, and should be read as the practical facial-oral disagreement relevant to screening deployment rather than a pure estimate of facial device error.

To assess the robustness of the group-level findings, a Monte Carlo sensitivity analysis added independent Gaussian noise to each participant’s value across 5,000 iterations at four perturbation levels (*σ* = 0.10, 0.15, 0.20, and 0.30 °C). Manufacturer accuracy specifications such as the device’s stated *±*0.2 °C are typically reported as a tolerance or maximum-error bound rather than a standard deviation, so the underlying noise distribution is not specified by the manufacturer. Testing *σ* from 0.10 °C to 0.30 °C brackets this ambiguity, spanning both a stricter reading of the spec as approximately one SD and a more conservative reading treating it as a wider bound. The ANOVA was refit at each level to evaluate the stability of *η*^2^ and of the group-mean ordering, the latter defined as the fraction of iterations in which the rank order of all five group means matched the unperturbed data.

## 4 Results

### 4.1 Analytic Cohort

The final cohort comprised 951 adults aged 18–30, of whom 40% were male. White participants formed the largest group (*n* = 476, 50.1%), followed by Asian (*n* = 244, 25.6%), Black or African-American (*n* = 132, 13.9%), Hispanic/Latino (*n* = 53, 5.6%), and Multiracial (*n* = 46, 4.8%). Mean *T*_CEmax_ ranged from 35.60 °C to 35.88 °C across the five categories, and mean *T*_max_ from 35.96 °C to 36.19 °C. Full group-level descriptive statistics are given in Table 1.

**Table 1:** Descriptive statistics for the final analytic cohort (*N* = 951). Values are mean *±* standard deviation (SD).

| Race and Ethnicity | $n$ | Male (%) | Age (y)<sup>a)</sup> | $T_{\text{CEmax}}$ ($^\circ\text{C}$) | $T_{\text{max}}$ ($^\circ\text{C}$) |
| --- | --- | --- | --- | --- | --- |
| Asian | 244 | 41 | $21.63 \pm 2.85$ | $35.60 \pm 0.59$ | $35.96 \pm 0.55$ |
| Black or African-American | 132 | 41 | $21.30 \pm 2.66$ | $35.63 \pm 0.66$ | $36.03 \pm 0.61$ |
| Hispanic/Latino | 53 | 38 | $20.92 \pm 2.41$ | $35.80 \pm 0.49$ | $36.13 \pm 0.54$ |
| Multiracial | 46 | 39 | $21.00 \pm 2.84$ | $35.81 \pm 0.70$ | $36.14 \pm 0.64$ |
| White | 476 | 40 | $20.94 \pm 2.64$ | $35.88 \pm 0.62$ | $36.19 \pm 0.59$ |

a) Age was recorded in categorical bins of unequal width (18–20, 21–25, 26–30, and an overlapping 21–30 category); continuous age was not available, so reported summaries are midpoint-based approximations. Overall: midpoint-based age $21.17 \pm 2.70$ years; 40% male.

### 4.2 Group Differences

Table 2 reports the full set of statistical tests. One-way ANOVA showed statistically significant effects of race and ethnicity on both measures, though the total spread between group means was only 0.2 °C to 0.3 °C (Figure 1), with a consistent ordering from lowest (Asian) to highest (White) across both outcomes.

**Table 2:** Statistical tests for the effect of race, ethnicity, and gender on facial temperature.

| Test | Statistic | $T_{CEmax}$ | $T_{max}$ |
| --- | --- | --- | --- |
| One-way ANOVA, ethnicity | $F(4, 946)$ | 9.95 | 6.73 |
| | $p$ | $< 0.001$ | $< 0.001$ |
| | $\eta^2$ | 0.04 | 0.03 |
| One-way ANOVA, gender | $F(1, 949)$ | 28.70 | 6.46 |
| | $p$ | $< 0.001$ | 0.011 |
| | $\eta^2$ | 0.03 | 0.007 |
| Brown–Forsythe | $F(4, 946)$ | 0.31 | 0.06 |
| | $p$ | 0.873 | 0.993 |
| Kruskal–Wallis | $\chi^2(4)$ | 52.54 | 36.20 |
| | $p$ | $< 0.001$ | $< 0.001$ |

**Figure 1:**
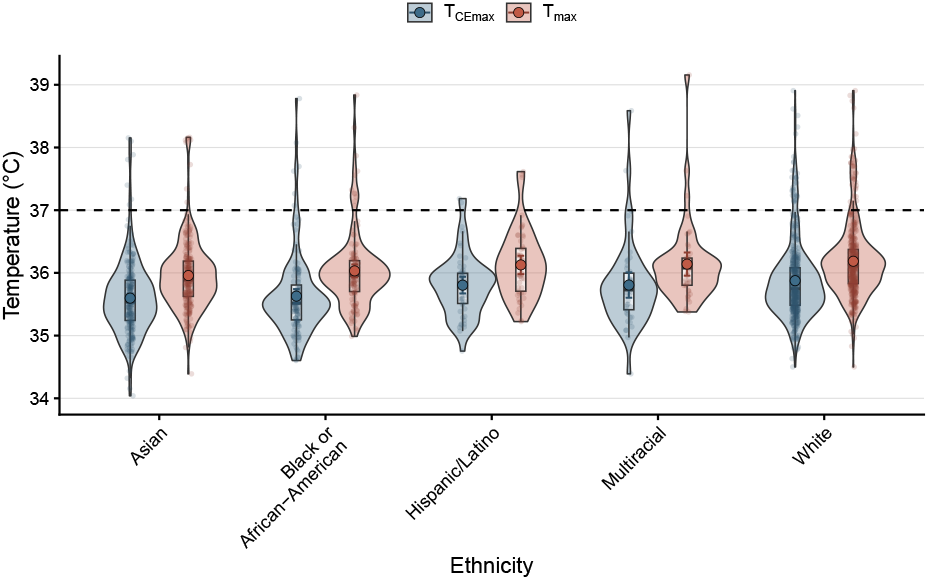
Distribution of participant-level mean *T*_CEmax_ and *T*_max_ by administratively encoded race and ethnicity (*N* = 951). Central markers indicate group means with 95% confidence intervals; dashed line indicates 37 °C as a historical reference value.

Primary hypotheses were evaluated at a conservative threshold of *α* = 0.001. Secondary covariate checks for gender yielded *F* (1, 949) = 28.70, *p <* 0.001, *η*^2^ = 0.03 for *T*_CEmax_ and *F* (1, 949) = 6.46, *p* = 0.011, *η*^2^ = 0.007 for *T*_max_. The latter did not meet the primary threshold, so it is treated as an exploratory covariate observation rather than a primary finding. Within the linear mixed-effects framework incorporating both demographic predictors and acquisition round, the fixed effect of race and ethnicity remained statistically significant for both measures.

Variance was homogeneous across groups (Brown–Forsythe), and the Kruskal–Wallis test confirmed the group effect was robust to the non-normality reported in Table 3, which gives per-group Shapiro–Wilk and outlier diagnostics.

**Table 3:** Per-group Shapiro–Wilk normality test and outlier check (Tukey IQR rule) for *T*_CEmax_ and *T*_max_.

| Ethnicity | $n$ | $W$ | $T_{\text{CEmax}}$ | | $W$ | $T_{\text{max}}$ | |
| --- | --- | --- | --- | --- | --- | --- | --- |
| | | | $p$ | Outliers (%) | | $p$ | Outliers (%) |
| Asian | 244 | 0.931 | < 0.001 | 4.1 | 0.918 | < 0.001 | 3.7 |
| Black or African-American | 132 | 0.849 | < 0.001 | 8.3 | 0.865 | < 0.001 | 8.3 |
| Hispanic/Latino | 53 | 0.963 | 0.099 | 7.5 | 0.945 | 0.016 | 5.7 |
| Multiracial | 46 | 0.862 | < 0.001 | 10.9 | 0.765 | < 0.001 | 8.7 |
| White | 476 | 0.894 | < 0.001 | 6.7 | 0.908 | < 0.001 | 7.4 |

Table 4 reports all ten pairwise comparisons across the five groups (Tukey HSD, 95% CI, adjusted *p*). For *T*_CEmax_, only White differed significantly from Asian and from Black or African-American; for *T*_max_, only White versus Asian reached significance. The overall ANOVA effect is therefore driven by a small number of group pairs rather than broad separation across all five categories.

**Table 4:**
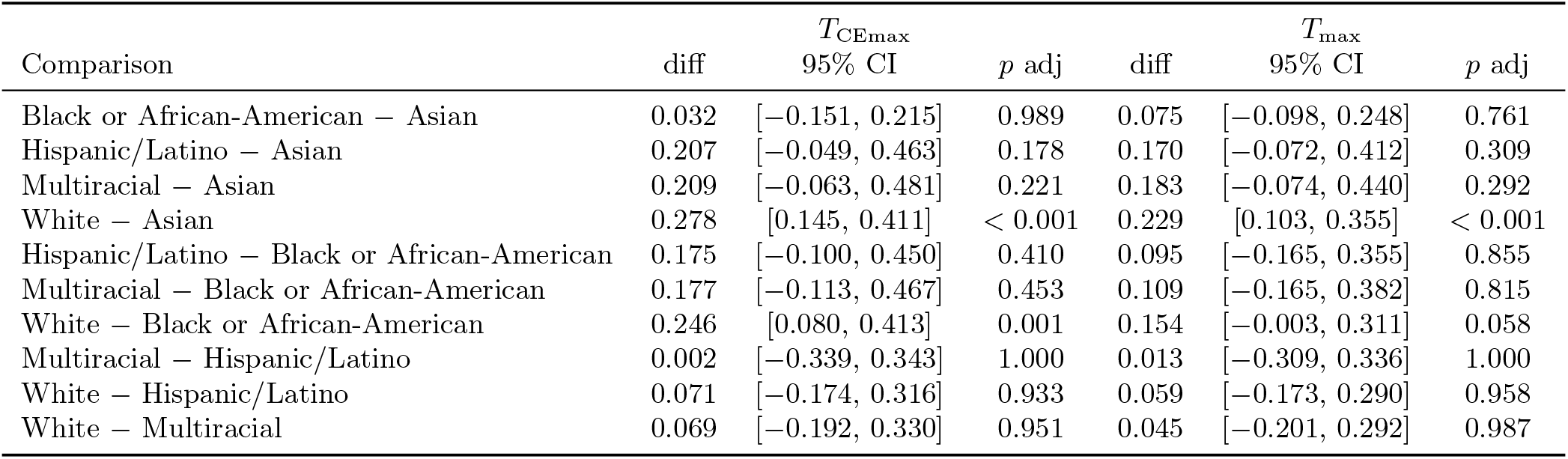
Pairwise ethnicity comparisons (Tukey HSD, 95% CI) for *T*_CEmax_ and *T*_max_.

| Comparison | diff | $T_{\text{CEmax}}$ | $p$ adj | diff | $T_{\text{max}}$ | $p$ adj |
| --- | --- | --- | --- | --- | --- | --- |
|  |  | 95% CI |  |  | 95% CI |  |
| Black or African-American – Asian | 0.032 | [−0.151, 0.215] | 0.989 | 0.075 | [−0.098, 0.248] | 0.761 |
| Hispanic/Latino – Asian | 0.207 | [−0.049, 0.463] | 0.178 | 0.170 | [−0.072, 0.412] | 0.309 |
| Multiracial – Asian | 0.209 | [−0.063, 0.481] | 0.221 | 0.183 | [−0.074, 0.440] | 0.292 |
| White – Asian | 0.278 | [0.145, 0.411] | < 0.001 | 0.229 | [0.103, 0.355] | < 0.001 |
| Hispanic/Latino – Black or African-American | 0.175 | [−0.100, 0.450] | 0.410 | 0.095 | [−0.165, 0.355] | 0.855 |
| Multiracial – Black or African-American | 0.177 | [−0.113, 0.467] | 0.453 | 0.109 | [−0.165, 0.382] | 0.815 |
| White – Black or African-American | 0.246 | [0.080, 0.413] | 0.001 | 0.154 | [−0.003, 0.311] | 0.058 |
| Multiracial – Hispanic/Latino | 0.002 | [−0.339, 0.343] | 1.000 | 0.013 | [−0.309, 0.336] | 1.000 |
| White – Hispanic/Latino | 0.071 | [−0.174, 0.316] | 0.933 | 0.059 | [−0.173, 0.290] | 0.958 |
| White – Multiracial | 0.069 | [−0.192, 0.330] | 0.951 | 0.045 | [−0.201, 0.292] | 0.987 |

### 4.3 Sensitivity Analysis

The Monte Carlo analysis perturbed each value with added Gaussian noise (Figure 2). Table 5 reports results across all four perturbation levels. Median *η*^2^ declined and the percentile range widened as *σ* rose from 0.10 °C to 0.30 °C for both measures. Group-mean ordering stability, the fraction of iterations preserving the complete rank order of the five group means, fell steadily over the same range, dropping below half for *T*_CEmax_ almost immediately and for *T*_max_ only at the highest perturbation level tested.

**Table 5:**
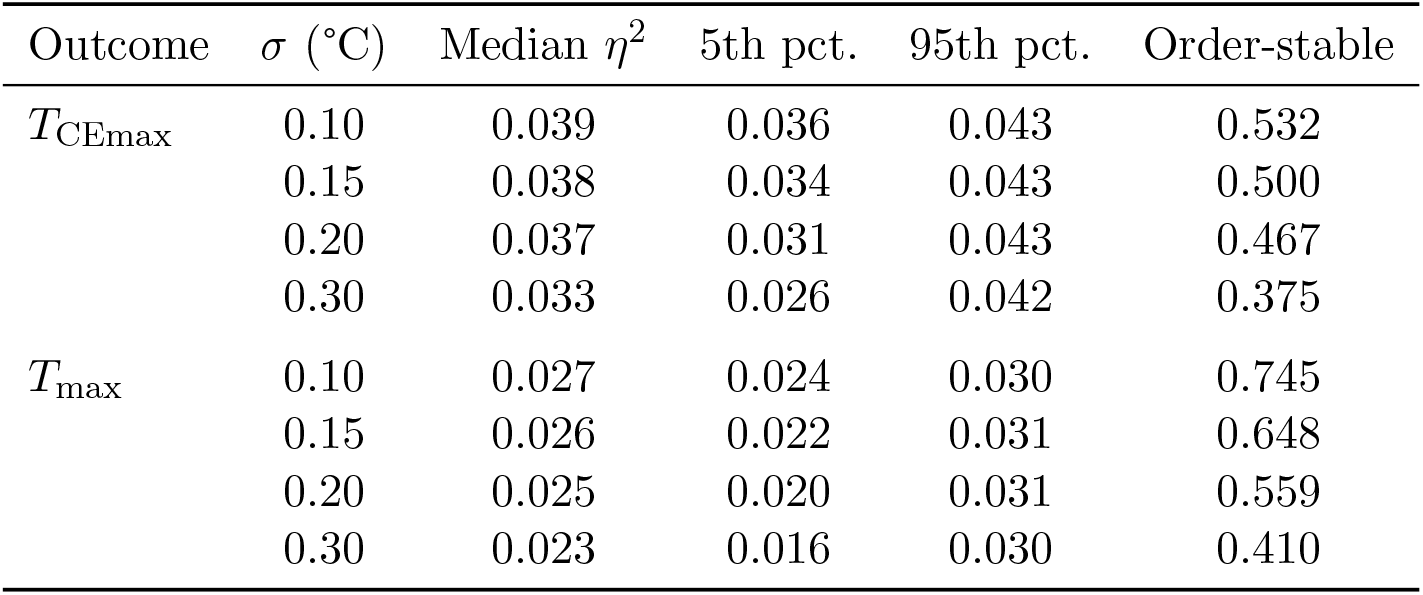
Monte Carlo sensitivity of ANOVA results to additive Gaussian measurement noise (SD = *σ*; 5000 iterations per level). Median *η*^2^ is reported with the 5th and 95th percentile range across iterations. Order-stable is the fraction of iterations preserving the complete rank order of the five group means.

| Outcome | $\sigma$ ($^{\circ}\text{C}$) | Median $\eta^2$ | 5th pct. | 95th pct. | Order-stable |
| --- | --- | --- | --- | --- | --- |
| $T_{\text{CEmax}}$ | 0.10 | 0.039 | 0.036 | 0.043 | 0.532 |
|  | 0.15 | 0.038 | 0.034 | 0.043 | 0.500 |
|  | 0.20 | 0.037 | 0.031 | 0.043 | 0.467 |
|  | 0.30 | 0.033 | 0.026 | 0.042 | 0.375 |
| $T_{\text{max}}$ | 0.10 | 0.027 | 0.024 | 0.030 | 0.745 |
|  | 0.15 | 0.026 | 0.022 | 0.031 | 0.648 |
|  | 0.20 | 0.025 | 0.020 | 0.031 | 0.559 |
|  | 0.30 | 0.023 | 0.016 | 0.030 | 0.410 |

**Figure 2:**
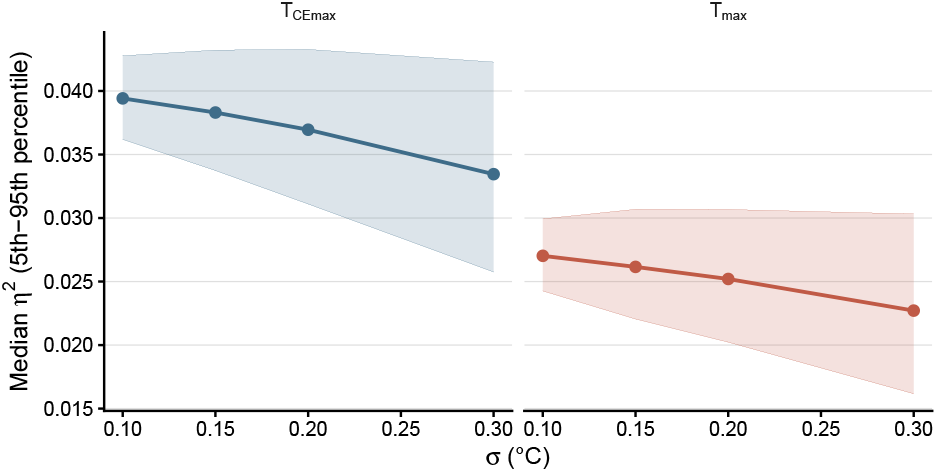
Monte Carlo sensitivity of *η*^2^ to additive Gaussian perturbations (*σ* =0.10 °C to 0.30 °C; 5000 simulations per level). Points represent median *η*^2^; shaded regions denote the 5th-95th percentile range. Full values are reported in Table 5.

### 4.4 Full Model Results

Table 6 reports the complete mixed-effects model coefficients for both outcomes. Ethnicity and gender remained significant fixed effects after accounting for repeated measures within participant, and acquisition round itself was a significant predictor, most strongly for *T*_max_, where the fourth round differed from the first by 0.83 °C.

**Table 6:** Linear mixed-effects model coefficients for *T*_CEmax_ and *T*_max_ (*temp ∼ Ethnicity* + *Gender* + *round* + (1|*SubjectID* )), with variance components and ICC.

| Fixed effect | $T_{\text{CEmax}}$ | | $T_{\text{max}}$ | |
| --- | --- | --- | --- | --- |
| | Estimate | $p$ | Estimate | $p$ |
| Intercept (Asian, female, round 1) | 35.444 | < 0.001 | 35.637 | < 0.001 |
| Black or African-American | 0.033 | 0.621 | 0.067 | 0.278 |
| Hispanic/Latino | 0.219 | 0.018 | 0.204 | 0.020 |
| Multiracial | 0.212 | 0.031 | 0.164 | 0.076 |
| White | 0.281 | < 0.001 | 0.228 | < 0.001 |
| Male | 0.224 | < 0.001 | 0.101 | 0.008 |
| Round 2 | 0.057 | < 0.001 | 0.158 | < 0.001 |
| Round 3 | 0.097 | < 0.001 | 0.182 | < 0.001 |
| Round 4 | 0.100 | < 0.001 | 0.828 | < 0.001 |
| Ethnicity (Type III Wald $\chi^2$) | $\chi^2(4) = 42.29, p < 0.001$ | | $\chi^2(4) = 28.48, p < 0.001$ | |
| Between-subject variance (SD) | 0.365 (0.604) |  | 0.302 (0.550) |  |
| Residual variance (SD) | 0.018 (0.136) |  | 0.092 (0.303) |  |
| ICC | 0.952 |  | 0.767 |  |

Table 7 reports Bland–Altman bias and limits of agreement against oral reference by ethnicity. Facial-oral bias itself differed significantly across groups for both measures, with Black or African-American participants showing the largest negative bias and White participants the smallest.

**Table 7:**
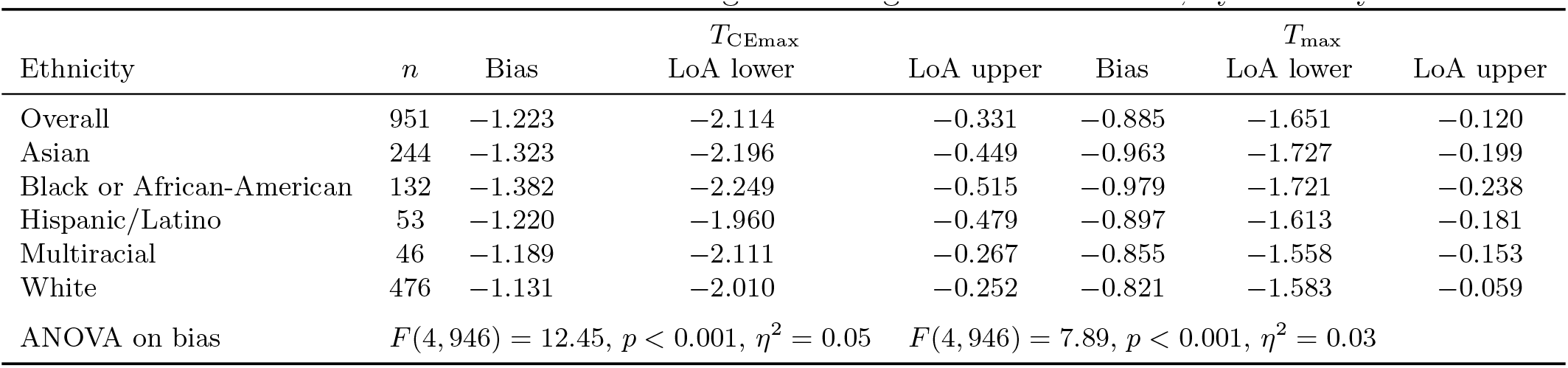
Bland–Altman bias and 95% limits of agreement against oral reference, by ethnicity.

The repeatability estimates for *T*_max_ diverge more than those for *T*_CEmax_ because acquisition round has a much larger systematic effect on *T*_max_. The pooled round-level SD (0.14 °C for *T*_CEmax_, 0.47 °C for *T*_max_) treats all round-to-round variation as noise. The mixed model instead includes round as a fixed effect, and for *T*_max_ the fourth round differs from the first by 0.83 °C

(Table 6), a drift plausibly driven by facial warming or cumulative exposure across a session. Once this systematic round effect is removed, the residual within-subject SD for *T*_max_ falls to 0.303 °C. *T*_CEmax_ shows much smaller round effects (0.06 °C to 0.10 °C), so its residual SD (0.136 °C) is close to the raw pooled estimate.

## 5 Discussion

### 5.1 Sources of Variance

The differences observed across race and ethnicity categories sit within a margin the instrument may not reliably resolve from measurement noise. Individual variation accounts for 95.2% of *T*_CEmax_ variance, leaving little variance for demographic categories to explain. Furthermore, because the total spread between the highest and lowest group means is only 0.2 °C to 0.3 °C, this gradient matches the scale of the measurement’s within-subject repeatability (0.136 °C) and the baseline device accuracy (*±*0.2 ^*°*^C), meaning the group-level signal is effectively buried within normal baseline noise.

Systematic variation across categories is present before any group comparison is made, with the facial–oral bias varying across categories by 0.251 °C. The Monte Carlo sensitivity analysis preserves the exact rank ordering of categories in only 53.2% of iterations for *T*_CEmax_ at the lowest noise level tested. In each iteration, Gaussian noise was added to each participant’s temperature, after which group means were recalculated and ranked from lowest to highest. A rank order was counted as preserved only if all five groups landed back in their original sequence. That ordering held in barely more than half of iterations even at this smallest perturbation, meaning noise consistent with the device’s own accuracy was enough to reorder the groups nearly as often as it left them intact. This is a direct consequence of group means clustered tightly enough that measurement noise reshuffles their order. The resulting ranking should not be interpreted as evidence of a stable physiological ordering, indicating that administrative labels impose discrete structure on a signal the instrument may not reliably resolve at this scale [4, 18].

### 5.2 Clinical Implications

The practical value of any fever screening system depends on whether the measurement it produces is precise enough to distinguish a febrile individual from a healthy one, and the agreement between facial and oral temperature in this dataset places that precision in question. The 95% limits of agreement for *T*_CEmax_ span 1.783 °C, meaning that for any given facial reading the corresponding oral temperature could fall anywhere within a range nearly two degrees wide, broad enough to place an individual on either side of the 38 °C fever boundary from the same measurement. These limits were measured under carefully controlled acquisition conditions and will only widen in real screening environments where imaging distance, head angle, ambient temperature, and patient acclimation vary. The between-group temperature differences identified across demographic categories span 0.2 °C to 0.3 °C, roughly one tenth to one sixth the width of the limits of agreement. Encoding demographic corrections at that scale should not be done, as it risks formalizing a measurement artifact as a biological difference and it could become embedded into the decision rules applied to individuals while leaving the underlying classification uncertainty unaddressed.

### 5.3 Downstream Propagation

The artifacts that undermine demographic thresholds also compromise the health informatics systems built on these data. When predictive models are trained on demographically labeled thermal inputs, they inadvertently learn these instrument-level differences as if they reflect true human physiology. These variations are then formalized within the model weights and decision rules. Because these automated systems treat administrative categories as biological facts, they risk encoding superficial sensor bias into downstream clinical models [4, 5].

### 5.4 Limitations

This study has several limitations. First, neither the IR camera nor the oral thermometer had verifiable calibration for this specific dataset. The source study paired each imaging session with an external blackbody reference for drift compensation [19], but no calibration schedule or drift record was released alongside the data. Device accuracy is therefore taken from the manufacturer specification and the source study’s own reported system-level accuracy [19–21]. It was not independently confirmed. If true device accuracy is worse than these stated values, a possibility supported by prior work showing infrared systems of this class often underperform their specification [3, 16], the group-level differences examined in this paper would fall even further inside the range of measurement noise. Second, skin pigmentation could not be evaluated as a distinct mechanism, separate from the broader optical effects, because the dataset records no objective pigmentation measure such as the Monk Skin Tone or Fitzpatrick Scale. As a secondary analysis of a public, de-identified dataset, this measure cannot be added retroactively; its absence limits this study to inferring pigmentation’s role indirectly, through its correlation with the administrative ethnicity categories, rather than testing it directly. Third, because the repeated measures were acquired within a single session among adults aged 18–30 under controlled indoor conditions, these data cannot establish whether individual baselines hold across days, seasons, or broader clinical populations.

## 6 Conclusion

Beyond evaluating a specific dataset, this study’s primary contribution is demonstrating a generalizable methodological check for health informatics when evaluating demographically stratified data: testing whether a statistically significant group difference (*p <* 0.001) across race and ethnicity categories survives perturbation at the scale of the instrument’s own accuracy. The observed significance in this cohort likely reflects the accumulation of individual physiology, instrument uncertainty, skin pigmentation and other optical effects, and administrative encoding within the same analytic pipeline. Individual variation accounts for over 95% of *T*_CEmax_ variance, the total spread between group means (0.2 °C to 0.3 °C) falls within the device’s nominal accuracy and an individual’s own measurement variability, and group-mean rankings for *T*_CEmax_ were preserved in only 53% of Monte Carlo iterations at the lowest perturbation level, collapsing further as noise approached the device’s accuracy limit.

When these artifacts are encoded into health informatics infrastructure without a record of their origin, clinical decision support systems, risk stratification tools, and population health dashboards inherit them as though they reflect human biology. Future work should establish instrumentation metadata standards capturing calibration state, acquisition conditions, and objective pigmentation measures, alongside longitudinal designs that follow individual baselines across days and environmental conditions. These recommendations are not unique to thermometry, as any analytic pipeline that treats administrative categories as biological inputs may reproduce this problem regardless of the instrument.

## Data Availability

The data analyzed in this study are publicly available through PhysioNet: Wang, Quanzeng, et al. "Facial and oral temperature data from a large set of human subject volunteers" (version 1.0.0). PhysioNet (2023). https://doi.org/10.13026/3bhc-9065

https://physionet.org/content/face-oral-temp-data/1.0.0/

## Acknowledgements

This research received no external funding. This study is a secondary analysis of publicly available, de-identified data. The original data collection was conducted under Institutional Review Board and Food and Drug Administration-related study governance [19, 20]. As this is a secondary analysis of de-identified data, no additional ethics approval was required. The thermographic dataset analyzed in this study is publicly available from PhysioNet [20]:https://physionet.org/content/face-oral-temp-data/1.0.0/

## References

[1] C. A. Wunderlich, Medical Thermometry and Human Temperature. The New York Printing Company, 1871. Abridged by E. Seguin.

[2] W. Chen, “Thermometry and interpretation of body temperature,” Biomedical Engineering Letters, vol. 9, pp. 3–17, 2019.

[3] N. Shinsuphan, P. Aphinan, T. Leeudomwong, and T. Sanponpute, “Assessment of the accuracy and reliability of clinical thermometers for body temperature measurements at the hospital: A laboratory study,” Measurement Science and Technology, vol. 35, no. 11, p. 115007, 2024. Article number 115007.

[4] P. Sankar, M. K. Cho, and J. Mountain, “Race and ethnicity in genetic research,” American Journal of Medical Genetics Part A, vol. 143A, no. 9, pp. 961–970, 2007.

[5] S. E. Ali-Khan, T. Krakowski, R. Tahir, and A. S. Daar, “The use of race, ethnicity and ancestry in human genetic research,” The HUGO Journal, vol. 5, no. 1–4, pp. 47–63, 2011.

[6] S. Taylor, “The origin of the thermometer,” Annals of Science, vol. 5, no. 2, pp. 129–156, 1942.

[7] D. Livingstone, Missionary Travels and Researches in South Africa. J. Murray, London, 1 ed., 1857. Smithsonian Libraries digitization.

[8] M. Sund-Levander and E. Grodzinsky, “Time for a change to assess and evaluate body temperature in clinical practice,” International Journal of Nursing Practice, vol. 15, no. 4, pp. 241–249, 2009.

[9] Z. Obermeyer, J. K. Samra, and S. Mullainathan, “Individual differences in normal body temperature: Longitudinal big data analysis of patient records,” BMJ, vol. 359, p. j5468, 2017. Article ID j5468.

[10] J. Waalen and J. N. Buxbaum, “Is older colder or colder older? the association of age with body temperature in 18,630 individuals,” The Journals of Gerontology Series A: Biological Sciences and Medical Sciences, vol. 66A, no. 5, pp. 487–492, 2011.

[11] J. Raiko, K. Koskensalo, and T. Sainio, “Imaging-based internal body temperature measurements: The journal Temperature toolbox,” Temperature, vol. 7, no. 4, pp. 363–388, 2020.

[12] M. Charlton, S. A. Stanley, Z. Whitman, V. Wenn, T. J. Coats, M. Sims, and J. P. Thompson, “The effect of constitutive pigmentation on the measured emissivity of human skin,” PLOS ONE, vol. 15, no. 11, p. e0241843, 2020. Article e0241843.

[13] S. E. Sonenblum, K. Jordan, G. T. John, A. Chung, M. Asare-Baiden, J. Pelkmans, J. W. Gichoya, V. S. Hertzberg, and J. C. Ho, “Impact of skin tone, environmental, and technical factors on thermal imaging,” PLoS One, vol. 20, no. 9, p. e0325091, 2025.

[14] W. auf der Strasse, D. P. Campos, C.J.A. Mendonça, J. F. Soni, J. Mendes, and P. Nohama, “Forehead, temple and wrist temperature assessment of ethnic groups using infrared technology,” Medical Engineering and Physics, vol. 102, p. 103777, 2022. Article number 103777.

[15] P. P. Naik and S. N. Farrukh, “Influence of ethnicities and skin color variations in different populations: A review,” Skin Pharmacology and Physiology, vol. 35, no. 2, pp. 65–76, 2021.

[16] S. J. L. Sullivan, J. E. Rinaldi, P. Hariharan, J. P. Casamento, S. Baek, N. Seay, O. Vesnovsky, and L. D. T. Topoleski, “Clinical evaluation of non-contact infrared thermometers,” Scientific Reports, vol. 11, no. 1, p. 22079, 2021.

[17] G. B. Dell’Isola, E. Cosentini, L. Canale, G. Ficco, and M. Dell’Isola, “Noncontact body temperature measurement: Uncertainty evaluation and screening decision rule to prevent the spread of covid-19,” Sensors, vol. 21, no. 2, p. 346, 2021.

[18] N. Pearce, S. Foliaki, A. Sporle, and C. Cunningham, “Genetics, race, ethnicity, and health,” BMJ, vol. 328, no. 7447, pp. 1070–1072, 2004.

[19] Y. Zhou, P. Ghassemi, M. Chen, D. McBride, J. P. Casamento, T. J. Pfefer, and Q. Wang, “Clinical evaluation of fever-screening thermography: Impact of consensus guidelines and facial measurement location,” Journal of Biomedical Optics, vol. 25, Sept. 2020.

[20] Q. Wang, Y. Zhou, P. Ghassemi, M. Chen, J. P. Casamento, and D. McBride, “Facial and oral temperature data from a large set of human subject volunteers.” PhysioNet, 2023. Version 1.0.0. Published May 24, 2023.

[21] Infrared Cameras Inc. (ICI), “8640 P-Series IR Camera: Data Specifications.” https://pim.galco.com/Manufacturer/ICI\%20-\%20Infrared\%20Cameras\%20Inc/TechDocuments/Data\%20Sheet/ici-8640-p-series-ir-camera-data-specifications_dat.pdf, 2021. Revision 4.2021-001. NETD < 20 mK; accuracy ±1 ?C or ±1% of reading (standard mode).

[22] S. Heidari, T. F. Babor, P. De Castro, S. Tort, and M. Curno, “Sex and gender equity in research: rationale for the SAGER guidelines and recommended use,” Research Integrity and Peer Review, vol. 1, no. 1, p. 2, 2016.

[23] J. M. Bland and D. G. Altman, “Statistical methods for assessing agreement between two methods of clinical measurement,” The Lancet, vol. 327, no. 8476, pp. 307–310, 1986.

